# A Retrospective Study to Determine the Impact of Psychedelic Therapy for Dimensional Measures of Wellness: A Qualitative Analysis of Response Data

**DOI:** 10.1101/2023.02.12.23285814

**Authors:** Victoria Di Virgilio, Amir Minerbi, Jenna Fletcher, Anthony Di Virgilio, Salena Aggerwal, Luke Sheen, Jagpaul Kaur Deol, Gaurav Gupta

## Abstract

**Background:** The World Health Organization (WHO) defines wellness as the optimal state of health of individuals and groups. No study to date has identified the impact of psychedelic medicines for optimizing wellness using a dimensional approach. Treatment effects can be measured more broadly using a composite score of participants’ global perceptions of change for pain, function, and mood scores. Given the precedence in previous work for retrospective study of participants’ self-medicating with these substances, the nature of this study design allows for a safe way to develop further evidence in this area of care, with wellness as the broad indication.

**Methods:** 65 civilian or military veterans between the ages of 18-99 self-identifying as having used psychedelic medicines for non-recreational therapeutic purposes in the last 3 years were recruited for this study. Participants completed various standardized questionnaires that will be analyzed in a separate study, while this study analyzed the qualitative experiences described in relation to the medicines used and coded them according to themes developed from review of previous literature.

**Results:** A total of 93 comments were analyzed. Participant comments were classified into these categories: mysticism and spirituality, functional improvement and self awareness, social connection and cultural impact, impact on medical and mental health conditions, neutral impressions, sensations or difficult experiences. Participants described impacts in these categories related to spiritual, physiological, psychological, and social improvements, as well as difficulties and complex emotions regarding the experience of using psychedelic medicines.

**Discussion:** Wellness of individuals or groups is not simply an absence of disease, symptoms, or impairments. Rather, it reflects the outcome of numerous personal characteristics, psychophysiology, and choices, expressed throughout one’s lifespan, unfolding in dynamic interaction with a complicated socio-cultural and physical environment. Participants that used psychedelic medicines described improvement of medical and mental health conditions, social interaction, spirituality, and overall function. In general, quality of life and wellness consequently improved after the use of these medicines based on established multidimensional factors.

**Conclusion:** The use of various psychedelic medicines appears to be associated with a broad range of qualitative experiences that could help clarify the mechanism of how they impact wellness in the future.

## Background

The World Health Organization (WHO) constitution states: “*Health is a state of complete physical, mental and social well-being and not merely the absence of disease or infirmity*.” An important implication of this definition is that health is more than merely the absence of disorders or disabilities. Health is a state of well-being in which an individual realizes their own abilities, can cope with the normal stresses of life, can work productively and is able to contribute to their community. (*Health and Wellbeing*, n.d.).

A study examining the WHOs factors of quality of life (QOL), which include physical health, mental health, emotional well-being, and social functioning, concluded that the QOL concept is multidimensional (Baernholdt et al., 2012). The WHO considers wellness as the realization of the full potential of an individual physically, psychologically, socially, spiritually, and economically (*Health Promotion Glossary Update*, n.d.), thus quality of life and wellness are congruent in some areas, specifically mental and physical health, and social functioning.

Multiple dimensions underlying health and wellness cut across the diagnostic groups employed in classical categorical systems. These criteria-based categorical diagnostic systems are convenient for providers and service-providers, and are thus still frequently employed in medicine, public policy, and industry (e.g. insurance). However, the current movement toward personalized, precision-based, and participant-centered healthcare has empowered individuals to pursue professional care from a wide array of medical and allied health service providers. This is irrespective of whether a particular individual is deemed to meet the criteria for a particular diagnosis (Dawson, 1989; Kraft & Goodell, 1993).

Consider for example that there is a significant comorbidity between mental health and chronic pain issues (Cucinello-Ragland & Edwards, 2021; Kleykamp et al., 2021; Kohrt et al., 2018), yet it is rare that participants afflicted with both are included in studies, therefore outcomes on a given treatment for either/both are challenging to differentiate (Kirsh, 2010; McParland et al., 2021; Myhr & Augestad, 2013; Weeks et al., 2016). Some authors argue a causative relationship, while others point to the overlap in symptomatology, functional issues, and treatments making diagnostic distinctions difficult (Aronoff, 2016; Fornasari, 2012; Kynast et al., 2013; Rieder et al., 2017).

Furthermore definitions employed by the Diagnostic and Statistical Manuals (DSM) show the interrater reliability is poor, diagnostic criteria lack specificity and treatment outcomes are not necessarily tied to clinical parameters (Bremmer et al., 2008; Hahn et al., 2010; Lacasse & Leo, 2005; Lieblich et al., 2015; Liu et al., 2015; Maas et al., 2009; Osimo et al., 2020). In addition, prescriptions for antidepressant medications have far outnumbered the amount of individuals diagnosed with depression, objective data supporting the DSMs diagnostic criteria for conditions is limited, and there have been concerns about industry influence on these guidelines in the past (Moncrieff, 2018; Whitaker, 2005). That is why numerous authors have advocated for a dimensional approach to evaluation which is based on symptomatology, severity, life impact and response to treatment, instead of diagnostic categories (Brown & Barlow, 2005; Coghill & Sonuga-Barke, 2012; Widiger & Samuel, 2005). This would allow for individuals with various degrees of symptomatology to be compared irrespective of their underlying clinical diagnosis.

There are numerous limitations to the existing treatments employed as standard of care for specific categorically defined conditions (Benish et al., 2008; Cloitre, 2009; Foa et al., 2009; Hembree et al., 2003; Karlin et al., 2010; Nemeroff et al., 2006; Rothbaum et al., 2006; *Treatment of Posttraumatic Stress Disorder*, 2008; Ursano et al., 2004). Moreover, various other studies have investigated aspects of wellness including risk factors, anthropomorphology, physical and emotional symptomatology, occupational impact, quality of life, societal participation and daily function (i.e. activities of daily living). Interventions aimed at improving wellness have included behavioral modifications, education, exercise programs, workplace modifications and preventative screening programs (Martínez-Lemos, 2015).

Emerging studies on the use of psychedelics for various indications have shown some early optimism in various categorical conditions, ranging from addiction, mood, pain and anxiety disorders, and end of life of care (Bornemann et al., 2021; Gasser et al., 2015; Malone et al., 2018; Nielson et al., 2018; Watts et al., 2017). Specifically, only categorical/diagnostic conditions like depression or pain have been used to select participants and categorize outcomes. This limits the ability to use the data in a real world setting where participants suffer from more than one health issue or do not meet any specific criteria for a condition. Also given that various psychedelic medicines function and are employed in a similar way, and are likely safe long term, it would be useful to comparatively study them for people self-medicating with psychedelics for their own wellness. This approach will hopefully help develop a better understanding of where these medicines provide relief and at what magnitude (Bornemann et al., 2021; Castellanos et al., 2020; Tupper et al., 2015).

Psychedelic drugs have varying pharmacological profiles that have strong effects on the conscious experience, and can be discussed in two classes. Classic psychedelics include lysergic acid diethylamide [LSD], psilocybin, dimethyltryptamine [DMT] and mescaline exert their effect through the serotonin system particularly the 5HT2A receptor. The other category are entactogens such as 3,4-methylenedioxy-methamphetamine (MDMA), which are primarily phenethylamines and amphetamines (Tupper et al., 2015). Emerging studies on the use of psychedelics for various indications have shown some early optimism in various categorical conditions, ranging from addiction, mood, pain and anxiety disorders, and end of life of care (Krebs & Johansen, 2013; Mayor, 2019; Schlag et al., 2021). Clinical evidence to date in the use of psychedelics for chronic pain is limited; however, several studies and reports over the past 50 years have shown potential analgesic benefit in cancer pain, phantom limb pain and cluster headaches (Castellanos et al., 2020).

There are numerous limitations of existing studies, with more data required to determine applications to clinical practice (Elsey, 2017; Herzog et al., 2018; Pratt et al., 2019; Tupper et al., 2015). However, possible clinical benefits include; reduced severity of anxiety/mood and possibly pain symptoms, longer term clinical effect without maintenance therapy, less reliance on plant-based cannabis and possible reduced rates of substance abuse and relapse, bolstered prosocial behavior, reduced public healthcare utilization and improved reintegration from a personal, community, occupational (Elsey, 2017; Herzog et al., 2018; Nielson et al., 2018; Schlag et al., 2021; Tupper et al., 2015).

Despite the early optimism much needs to be understood about the indications, contraindications, dosing, relative efficacy compared to other similar treatments, impact on dimensional aspects of wellness in “non pathologic” states, and cost benefit. No study to date has looked at the impact of psychedelics for treatment of wellness as specifically only categorical/diagnostic criteria have been used to select participants and categorize outcomes. This further limits the ability to extrapolate outcomes to a real world setting (Brown & Barlow, 2005; Widiger & Samuel, 2005).

Given the overlap in mechanism of action of the various psychedelics, the known long term safety profile and the similarity in proposed clinical protocols for use, a dimensional approach will yield a more robust understanding of where these medicines help and by how much. Treatment effects can therefore be measured more broadly using a composite score of participant global perception, along with pain, function and mood scores, which has not been done to date.

Quantitative outcomes are generally more efficient to look at, but may not fully reflect the complexity of an experience. Previous qualitative studies regarding the use of psychedelic therapy have used various coding strategies to develop themes and categories based on therapy session transcripts. These techniques include thematic analysis (Clarke & Braun, 2017; Watts et al., 2017), computer assisted qualitative data analysis software (Belser et al., 2017), and qualitative content analysis (Gasser et al., 2015). These studies found that LSD-assisted psychotherapy in participants with severe mental illness associated with a life threatening disease provided secure and stable treatment results along with no serious negative effects (Gasser et al., 2015). In addition, psilocybin-assisted psychotherapy may provide psychologically distressed cancer participants with an efficacious treatment for mental and emotional distress (Belser et al., 2017). The effects may be divergent to antidepressants and short term talk therapies (Watts et al., 2017). The methods for analyzing qualitative data must reflect a broad range of experiences from participants, thus a compilation of these methods allows for an in depth analysis.

The overall aim of this specific study is to retrospectively assess the participants’ qualitative experience of using psychedelic medicines for improving their own wellness. Quantitative results related to the impact on wellness will be presented elsewhere.

## Methods

Given the previous precedence for retrospective study of participants self-medicating with psychedelics, the nature of this study design allows for a safe way to develop further evidence in this area of care (Bornemann et al., 2021; Schindler et al., 2015). The clinical trial registration number is NCT05469243.

65 participants self-identifying as having used psychedelic medicine for non-recreational purposes were recruited for this study. To be considered eligible for admission to the study they were civilian or military veterans between the ages of 18-99 with a self-reported past of psychedelic medicine used for therapeutic purposes in the last 3 years. Past psychedelic medicine use for only non-therapeutic purposes (as determined by the participant) was excluded, as well as active military members.

In addition to demographic information, participants completed the following psychometric measures: Patient Global Impression of Change (PGIC) scale measured for overall quality of life, anxiety, mood, pain and disability subscales, the Pain, Enjoyment of Life and General Activity (PEG) scale, the Hospital Anxiety and Depression (HAD) scale, and the Disability Index (DI). Participants also provided information on their past medical history, nature/indications for use (ie. depression, PTSD, chronic pain, other), and adverse events. The scores are expressed on the basis of how they felt at the time of reporting in relation to the numerous dimensions of wellness. No change in scores were measured. A deeper quantitative analysis will be provided in a separate study.

This specific article will examine the qualitative responses of the data collected in the comments. Comments were classified into categories using a modification of the previously described qualitative paper coding techniques (Belser et al., 2017; Gasser et al., 2015; Watts et al., 2017) and thematic analysis (Clarke & Braun, 2017). The comments were then analyzed by integrating categories from other previous qualitative studies (Belser et al., 2017; Gasser et al., 2015; Watts et al., 2017). Thematic analysis included the categories developed for this paper based on previous work, in order to identify the main themes presented in the comments (Watts et al., 2017).

Two authors (VD, GG) organized and reviewed the comments using the primary medicine and themes derived, which are presented in Appendix A. The primary categories applied were A) Mysticism/spirituality; B) Functional improvement/self awareness; C) Social connection/cultural impact; D) Impact on medical/mental health condition; and E) Neutral impression, sensations or difficult experiences. Comments were also further sub-categorized using the same themes if required.

## Results

Of the 65 questionnaires completed 38 were by women, with an average age of 45.1 years old. Twenty of the 38 women were unemployed or on a leave of absence compared to only 5 men, while the most cited medical conditions were anxiety and depression. Psilocybin and Ayahuasca were the most common medicines used, while 11 participants did not report which psychedelic they had used most recently. Average scores for the participant reported outcomes fell into the mild-moderate range depending on the measure. Table 1 & 2 further describes the demographics and outcomes of the various participant reported outcomes.

**Table1:** Demographics of Participants.

|  | Men (n=24) | Women (n=38) | Other (n=2) |
| --- | --- | --- | --- |
| *Age* |  |  |  |
| Average | 45.4 | 45.1 | 59 |
| *Work* |  |  |  |
| Full-time | 17 | 13 | 1 |
| Part-time | 2 | 5 | 1 |
| Unemployed and/or current leave of absence | 5 | 20 | 0 |
| *Diagnoses Reported* |  |  |  |
| Addiction | 7 | 7 | 0 |
| Anxiety | 10 | 27 | 0 |
| Chronic pain | 8 | 18 | 1 |
| Depression | 15 | 25 | 1 |
| End of life distress | 0 | 2 | 0 |
| PTSD | 8 | 19 | 1 |
| *Most Recent Medication Used* |  |  |  |
| Ayahuasca | 2 | 6 | 0 |
| Ecstasy | 0 | 3 | 0 |
| Ibogaine | 1 | 1 | 0 |
| Ketamine | 1 | 4 | 0 |
| Lysergic Acid Diethylamide | 1 | 2 | 0 |
| Cannabis | 2 | 1 | 0 |
| Methamphetamine | 0 | 2 | 0 |
| Psilocybin | 12 | 13 | 2 |
| None | 5 | 6 | 0 |

**Table 2:** Pain, Enjoyment and General Activity Scale (PEG), Disability Index (DI), and Hospital Anxiety and Depression (HAD) scores, including HAD depression and anxiety subscales.

|  | Men (n=24) | Women (n=38) | Other (n=2) |
| --- | --- | --- | --- |
| *Current PEG score* |  |  |  |
| Average | 11.5 | 14.0 | 18.5 |
| *Current DI score* |  |  |  |
| Average | 23.8 | 32.0 | 37.5 |
| *Current HAD score* |  |  |  |
| Average | 15.2 | 17.9 | 20 |
| *Current HAD depression subscale* |  |  |  |
| Average | 6.4 | 7.1 | 8.5 |
| *Current HAD anxiety subscale* |  |  |  |
| Average | 8.8 | 10.8 | 11.5 |

Note: PEG (Pain, Enjoyment and General Activity Scale), DI (Disability Index), HAD (Hospital Anxiety and Depression Scale)

**Table 3:**
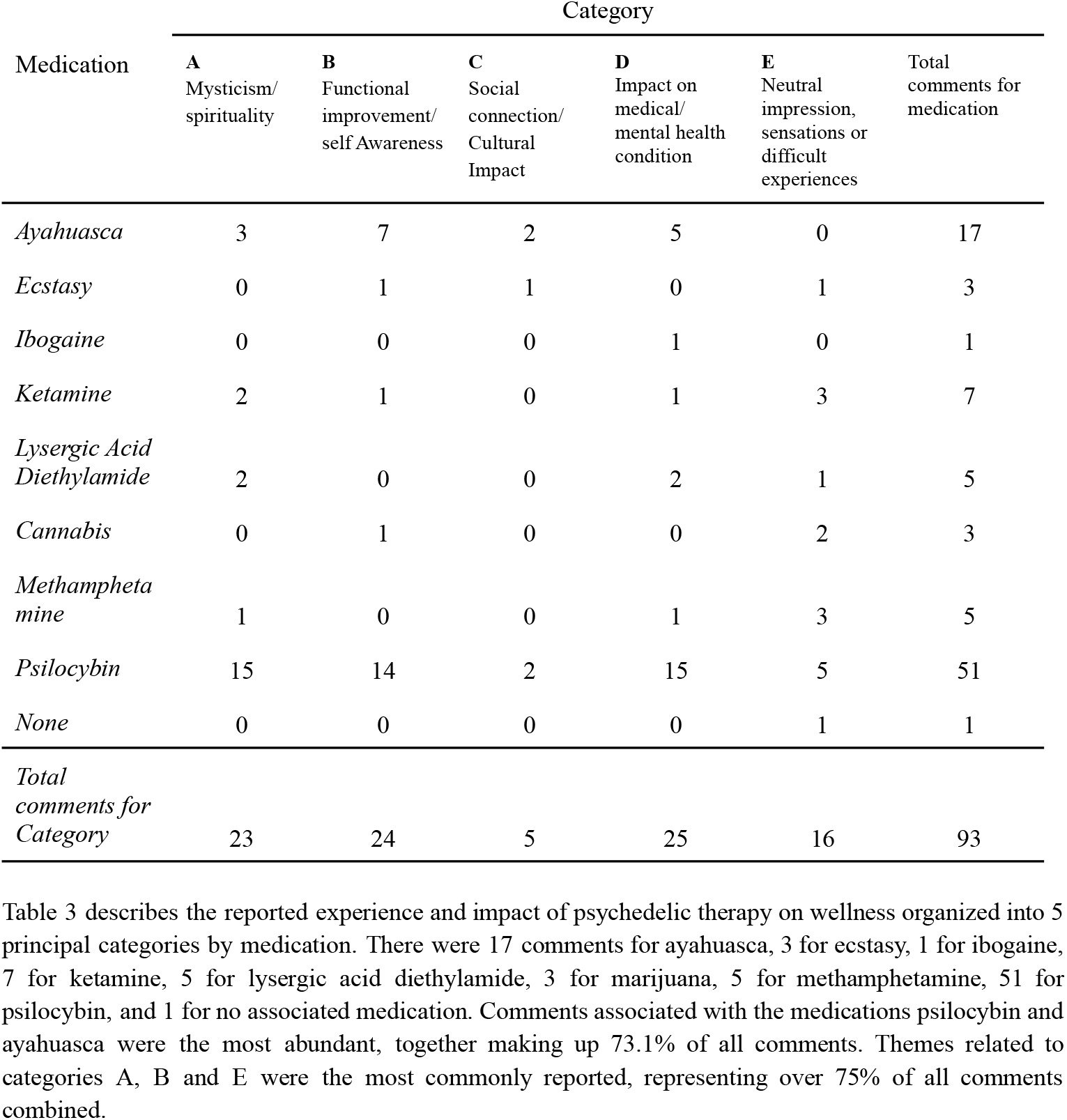
Medications taken resulting in comments from participants describing the impact and experience of psychedelic therapy for wellness associated with specific categories.

Numerous comments had multiple thematic elements. Table 4 shows the number of comments categorised and sub categorised into each theme. Three of the comments left by participants were originally in French, but were translated into English for the purposes of comprehension and consistency. No other changes were made to the comments. These are displayed in full in Appendix A.

**Table 4:** Qualitative analysis of comment occurrences in categories, including subcategory occurrences.

| Categories | Occurrences |  |  |  |
| --- | --- | --- | --- | --- |
|  | Category | Subcategory | Total | % |
| **A**<br>Mysticism/ spirituality | 23 | 13 | 36 | 38.7 |
| **B**<br>Functional improvement/ self Awareness | 24 | 11 | 35 | 37.6 |
| **C**<br>Social connection/ Cultural Impact | 5 | 9 | 14 | 15.1 |
| **D**<br>Impact on medical/ mental health condition | 25 | 10 | 35 | 37.6 |
| **E**<br>Neutral impression, sensations or difficult experiences | 16 | 23 | 39 | 41.9 |

93 comments total, Appendix A.

### Mysticism and Spirituality

Some participants described experiences and lasting effects of a religious or spiritual essence. They expressed that psychedelic therapy provided support for reconnection or revelational moments related to their connection with various religions, mind and body, soul, and life’s purpose. Comments ranged from mystical and spiritual experiences to ego dissolution, detachment, confusion. Subcategories from previous qualitative studies that were integrated into this classification included connection to a spiritual principle, and/or oneself (Watts et al., 2017). Spiritual and religious experiences were found in 36 comments, including comments from the subcategories.

> “shaking; nausea; yawning; visual effects (intensely detailed, exquisite and ethereal patterns and visions; connection with and guidance from holy spirit; revelatory messages of life’s purpose and meaning; unforgettable experiences of infinite, eternal, divine love”

This participant revealed how they felt connected to religion and spirituality. Their experience with psychedelic therapy offered insights to life’s meaning and allowed them to perceive lasting feelings of love.

### Functional Improvement and Self Awareness

Many of the participants expressed valuable improvement of their daily activities as well as newfound self-awareness leading to more positive daily experiences. Functional improvement has to do with the improvement of daily activities. Some of the comments discussed release of emotional loads, positive changes to activities and habits, and elements of self awareness. Subcategories that were integrated into this classification based on previous qualitative studies are revised life priorities (Belser et al., 2017), increased quality of life, faster progress, physical well-being, increased awareness, and important introspection (Gasser et al., 2015). Functional improvement and self awareness was found to have been improved in 35 comments, including comments classified in subcategories.

> “I have a clearer understanding of where my mental health and any physical pain play a role in my life; I have a better acceptance of things in my life and am not held back by these things.”

This participant developed stronger self awareness which allowed them to improve their daily functioning.

### Social Connection and Cultural Impact

Social connection necessary for interpersonal relationships was found to be improved in a number of participants as well as specific cultural experiences provided by the psychedelic medicine. A participant who saw cultural impacts described how the medicine changed their perspective on indigenous worldviews. Other participants described various elements in which their interpersonal relationships became improved after integrating the experience with the psychedelic medicine into their lives, but some participants traversed through difficult experiences in order to reach the improvement in their relationships. Themes from previous qualitative studies that were integrated into this classification are improved relationships after treatment, from separateness to interconnectedness (Belser et al., 2017), increased importance of family (Gasser et al., 2015), connection to the world, and connection to others (Watts et al., 2017). Social connection and cultural elements were improved in 14 comments, including comments classified in subcategories.

> “purging, shaking, crying, spiritual experiences (feeling of being loved, feeling of connection to humanity), intense emotional processing (processing of past attachment injuries, meaning making, self-compassion, moving through unfinished business in interpersonal relationships [parents, past romantic attachments]”

Interpersonal relationships with parents and past romantic companions were topics of distress for this participant, but the psychedelic therapy for wellness allowed them to transcend those difficulties and improve future interpersonal relationships.

### Impact on medical and mental health condition

Participants experienced a variety of changes to their conditions resulting in an improved quality of life and improved wellness. This classification could resonate with many participants as they all have some sort of medical or mental health condition that may cause certain limitations of function, pain, and psychological distress. There were many accounts of reduced pain (general, localized and chronic pain), reduced anxiety and/or reduced symptoms associated with specific mental health conditions. Some participants described that the reduction in symptoms did not last long, and difficulties with accessing psychedelics. The subcategories from previous qualitative studies that were integrated into this classification along with general reduction in psychological and physiological conditions are reduced anxiety and increased mental strength (Gasser et al., 2015). Impact on both medical and mental health conditions were improved in 35 comments, including comments classified in subcategories.

> “Mental resolution, removed brain fog, relieved pain in my joints and head, relieved anxiety for up to about a month”

One participant expressed the impact of psychedelic therapy on various aspects of their mental and physical health. They experienced reduced symptoms associated with their anxiety and physical pain. Although the participant did factor in how the relief only lasted for a certain period of time after taking the psychedelic.

### Neutral impression, sensations or difficult experiences

Some participants experienced different impacts regarding the impact of psychedelic therapy for wellness that do not fall into a category that represents a form of mental, physical, or functional improvement. Participants left comments regarding side effects, setbacks due to lack of medication access, and various sensations including purging, nausea, vomiting, and shaking. Themes from previous qualitative studies that were integrated into this category include surrender or “letting go” following transient psychological distress, continued struggle, catharsis of powerful emotion (Belser et al., 2017), disconnection, and the confrontation of traumatic memories from childhood (Watts et al., 2017). This section represents participants that generally experienced physiological sensations, or difficult experiences, although some comments are classified in other subcategories. These themes were found in 39 comments, including comments classified in subcategories.

> “the only associated symptom that I got after a “Trip” was fatigue, some expected anxiety prior too”

As described in the participant comment, they experienced some effects such as anxiety and fatigue.

## Discussion

Wellness of individuals or groups is not simply an absence of disease, symptoms, or impairments. It is rather the outcome of numerous personal characteristics, psychophysiology, and choices, expressed throughout one’s lifespan, unfolding in dynamic interaction with a complicated socio-cultural and physical environment. The accumulation of research in fields such as physiology, medicine, and neuroscience has highlighted that comorbidity is the norm, and thus there is limited external validity or clinical utility to studies that isolate small samples of individuals with “pure” diagnoses by excluding the majority of persons (Endresen & Wintz, 1988; Grasser & Craft, 1984; Magnavita & Garbarino, 2017; C. K. Ross et al., 1987).

Despite the ubiquitous nature of suffering and vast array of choices, the relative and/or synergistic benefit of commonly employed treatments for wellness such as rehabilitation, mental health support, medications, injections and surgery have never been convincingly elucidated (Frass et al., 2012; Giannitrapani et al., 2019; Herman et al., 2005; van der Watt et al., 2008). Studies are limited by heterogeneity of conditions, lack of standardization of measurement, small sample sizes, lack of statistical power, small effect sizes and questionable durability of responses, (P. T. Ross & Bibler Zaidi, 2019; Zwahlen et al., 2008). This limits our ability to draw generalizable conclusions resulting in (a) a limited understanding of real world/societal implications of treatments; (b) a limited ability to extrapolate to varied populations/edge cases; (c) poor control for placebo effects; (d) a limited understanding of synergies between different types of care; as well as (e) a need to evolve care pathways and test the validity of current diagnostic criteria/consensus statements regarding care (Frass et al., 2012; Herman et al., 2005; P. T. Ross & Bibler Zaidi, 2019; Zwahlen et al., 2008). However these treatments are routinely used, often at great cost and unknown risk, in caring for people in many use cases such as non-pathologic suffering/life concerns, primary/secondary mental health conditions, chronic pain and end of life concerns (van der Watt et al., 2008).

Health systems and employers are increasingly looking to wellness programs as a cost-containment strategy, given the disproportionate share of medical expenditures, and absenteeism with noncommunicable diseases. One systematic review that looked at interventions targeting one of the four primary modifiable behaviors for chronic disease: physical activity, healthy diet, tobacco use, and harmful consumption of alcohol, did not show wellness programs deliver a positive return on investment within the first few years of initiation (Baid et al., 2021). Conversely, despite the high degree of heterogeneity between the studies, limited effect size and risk bias potential in other studies, some research has shown improvements in wellness for participants with multiple sclerosis, employees reporting stress, and health professionals at risk of burnout (Morrison & MacKinnon, 2008; O’Reilly et al., 2018; Richardson, 2017; Russell et al., 2020; Slavin et al., 2014; Weiss et al., 2021).

With respect to the participants in this study, many described improvements in multiple domains of wellness associated with psychedelic medicines. Similar to other studies, this included effects on medical and mental health conditions, social interaction, spirituality, and functional improvement, and quality of life, all established multidimensional factors related to wellness (Baernholdt et al., 2012). While there were negative consequences reported as well, in general there were one or more life domains positively impacted from a qualitative standpoint. The factors driving these effects will require further exploration in order to explain the mechanism of change for dimensional measures of wellness.

While Psilocybin and Ayahuasca were most commonly reported in this study, the thematic qualitative experiences were not unique to any single medicine, nor did a single theme always emerge for any given participant. This is consistent with existing literature, although the thematic changes at a granular level and effect size for each medicine and related experiences will likely require more study (Gasser et al., 2015; Watts et al., 2017). The long term goal of this type of research would be to determine if the perceived benefits are class or medicine specific, and what factors about the participant selection and treatment can conceivably be changed to facilitate outcomes (Belser et al., 2017). If this can further be tied to the neurobiology of the medicine and patient, a set of truly bespoke treatment options can emerge (Nielson et al., 2018).

The language used to express the experience may also be context, medicine and patient dependent. Examining how the words used to describe and reflect on the experience are influenced by various factors like preparation, expectation, health issues, culture and intention will be important. Defining a common language or validated set of themes with which to draw from might help facilitate understanding of these medicines and help extrapolate their applications more broadly (Malone et al., 2018). The effect of this understanding will help further clarify the combined qualitative and quantitative effects of these medications.

Limitations of this study include the retrospective nature and sample size. The results were based on self report, while time since administration and change in patient reported outcomes were not collected or standardized. Also unlike other studies, the qualitative information was not gathered through in person interviews, and commenting was left to the discretion of individual participants. Future studies can look further at a more structured thematic assessment to validate the categories developed here and to see what other themes may emerge. Finally while the impact of each medication for various domains needs to be elucidated more fully, understanding the neurophysiological effect will be key in treatment planning and personalization of care.

## Conclusion

The use of various psychedelic medicines appears to be associated with a broad range of qualitative experiences that could help clarify the mechanism of how they impact wellness in the future. Broadly speaking various circumscribed themes emerge when individual experiences are described, spanning multiple domains. While this qualitative impact could be class or medicine specific, eventually with a validated set of language tools and an improved understanding of the neurobiology, qualitative and quantitative outcomes may become more predictable. This would be imperative in designing patient specific treatments using these medicines in the future.

## Data Availability

All data produced in the present work are contained in the manuscript

## Appendix A participant Comments

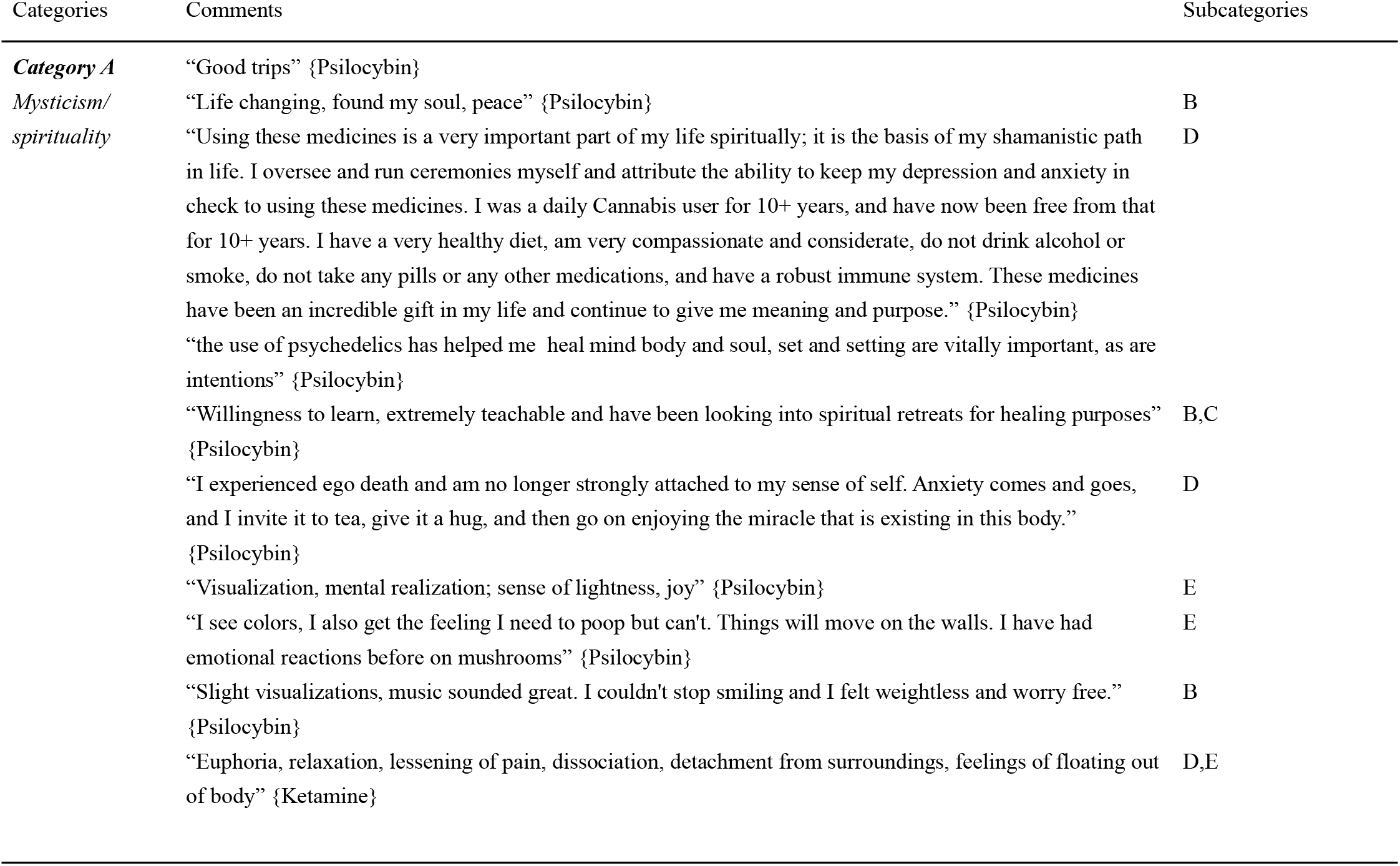
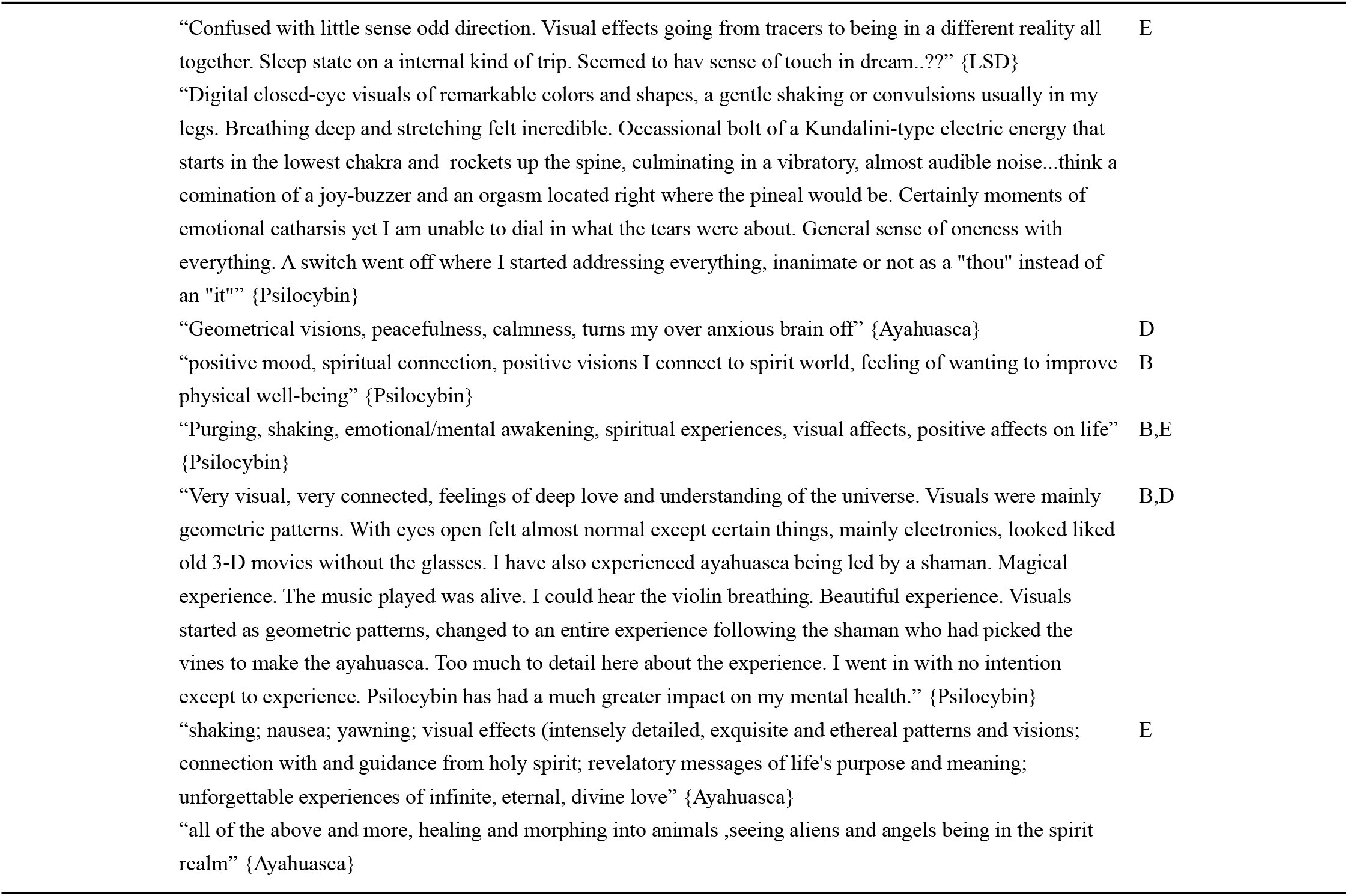
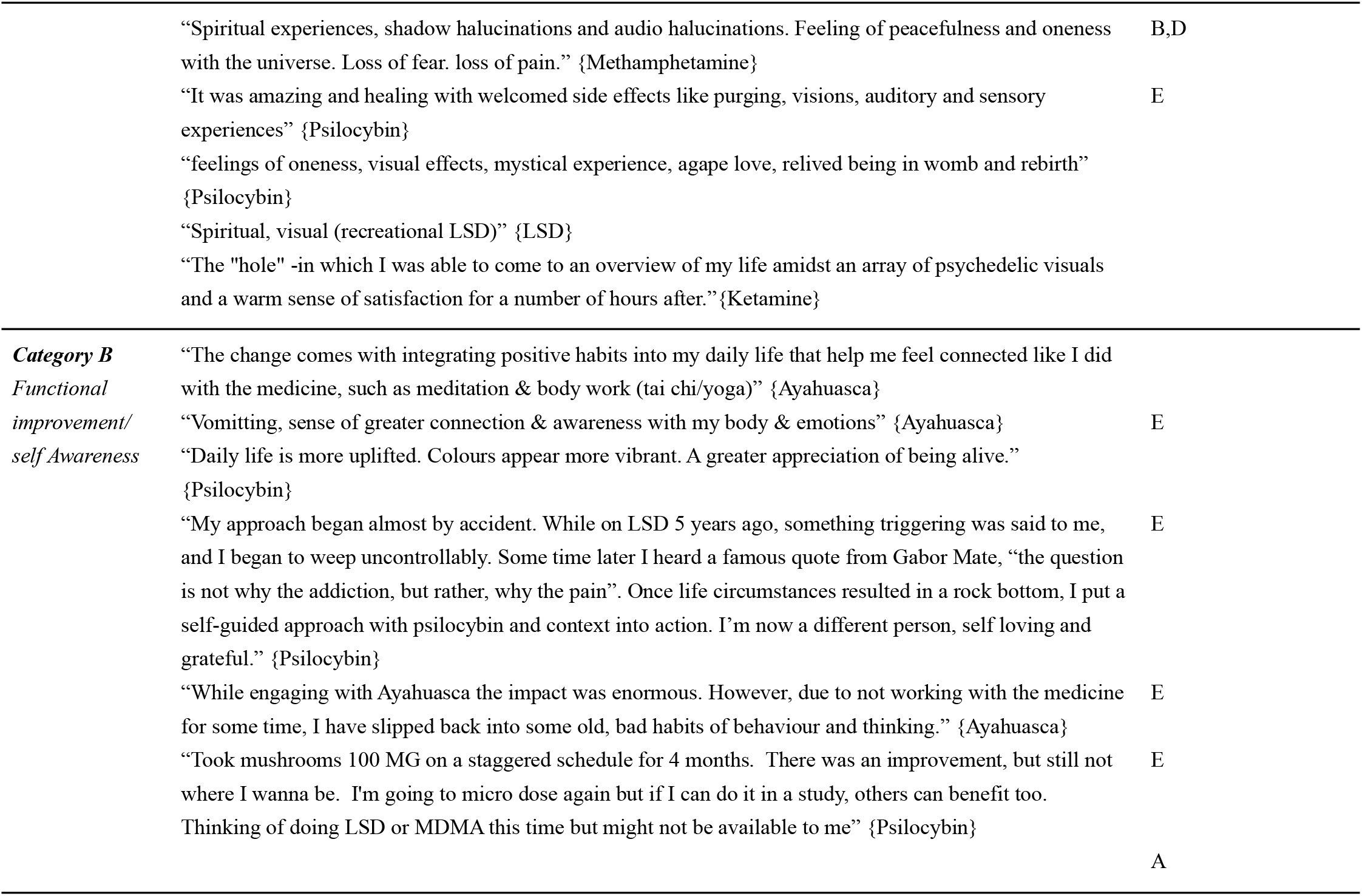
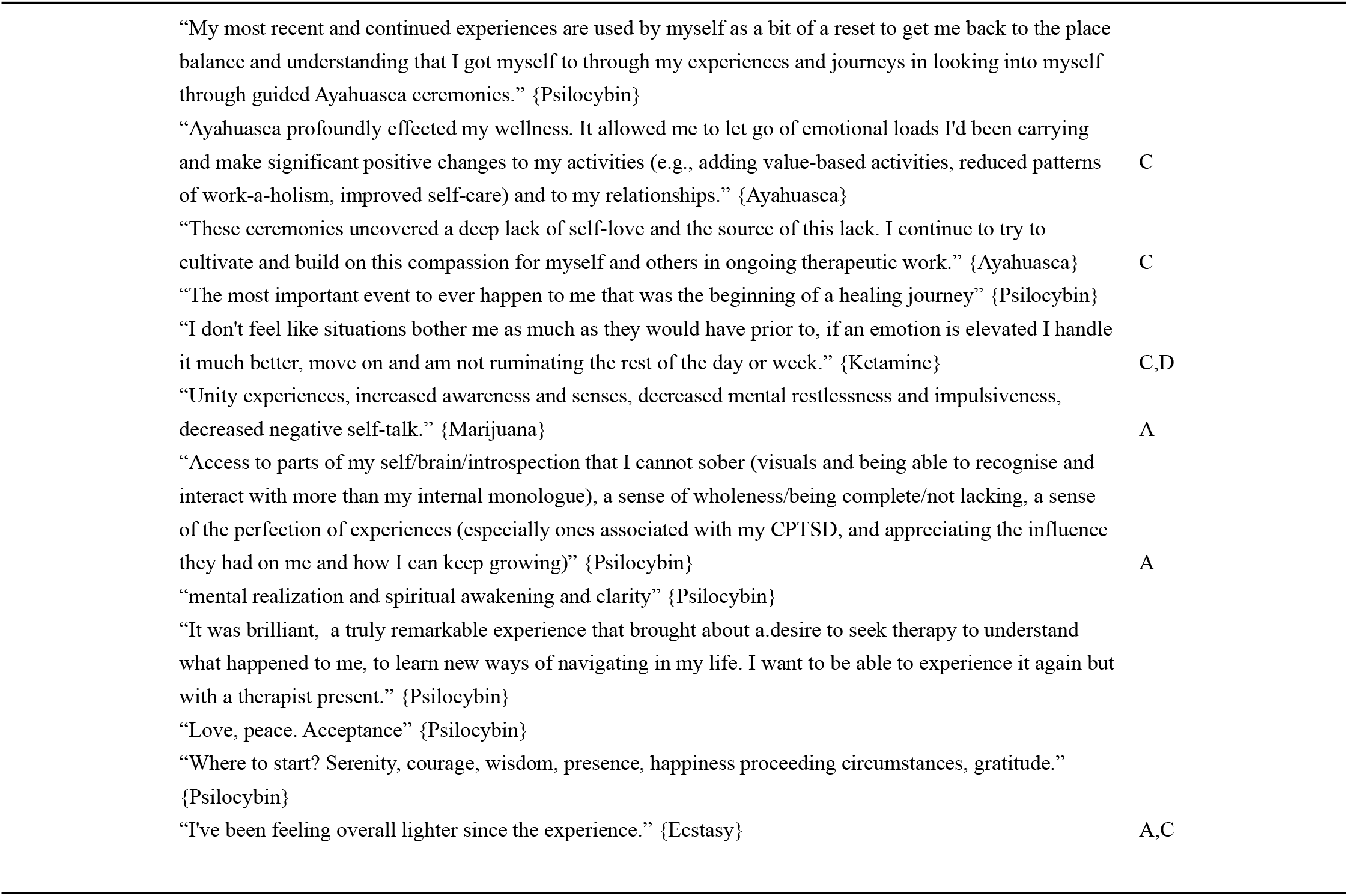
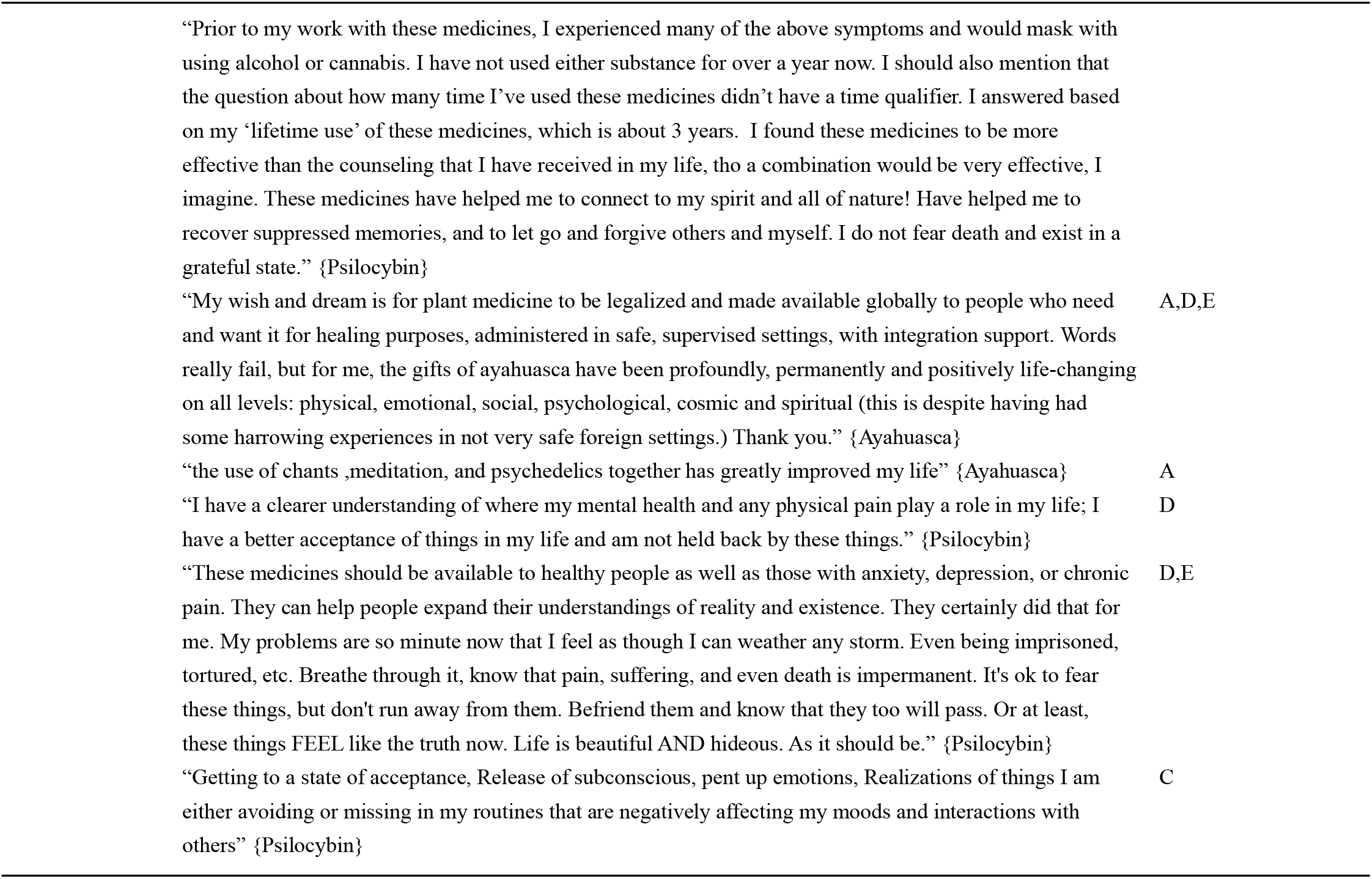
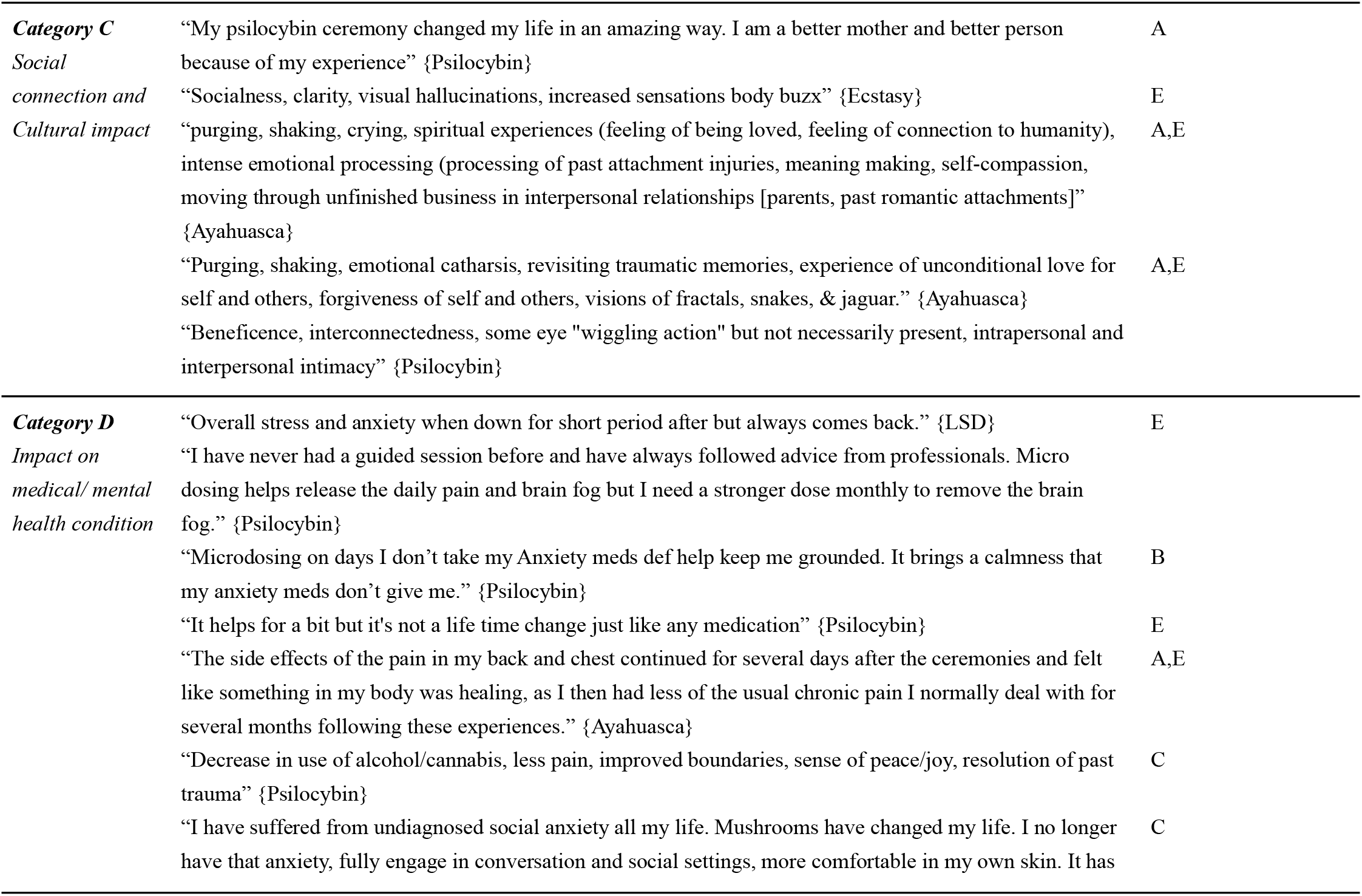
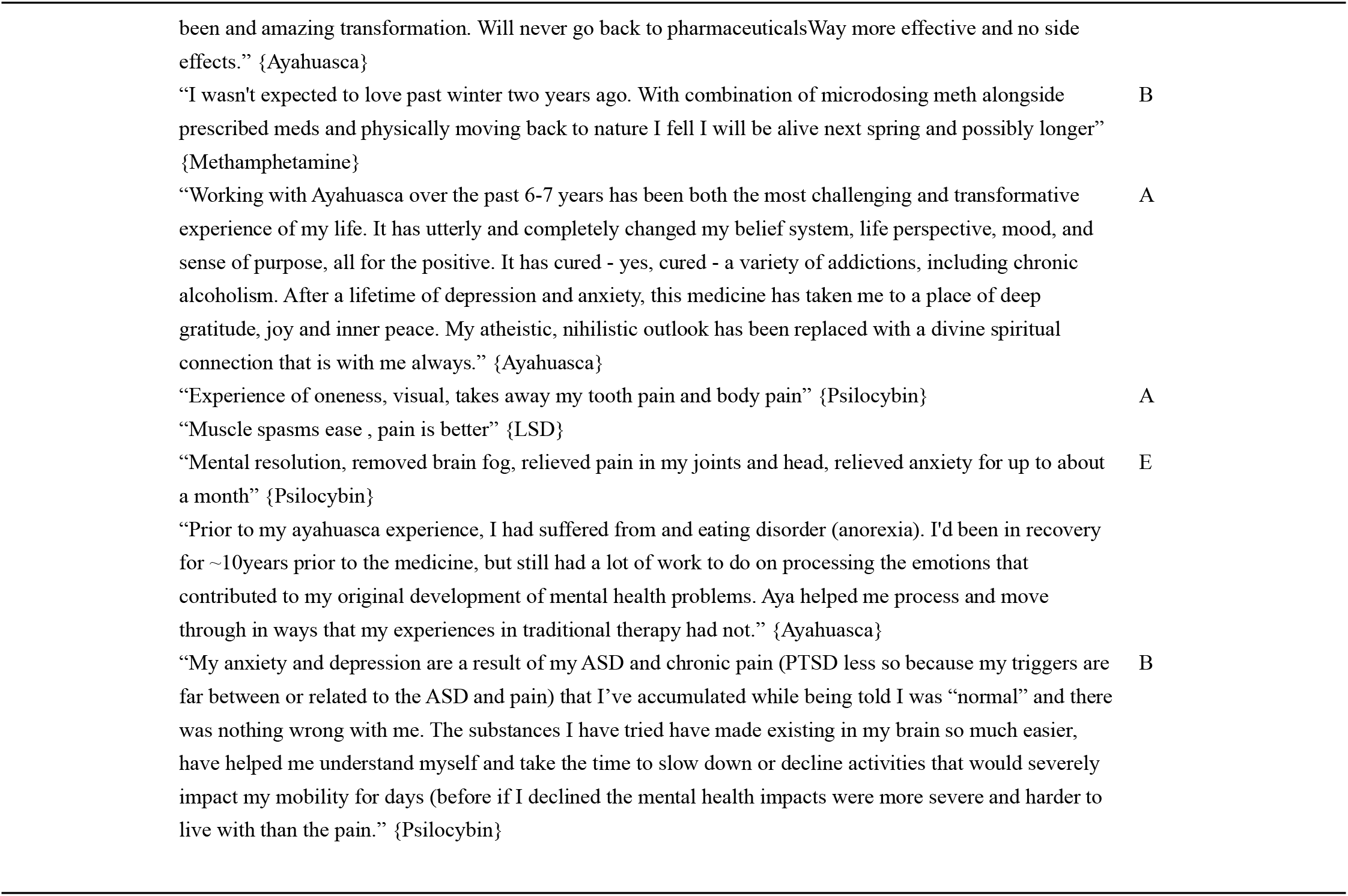
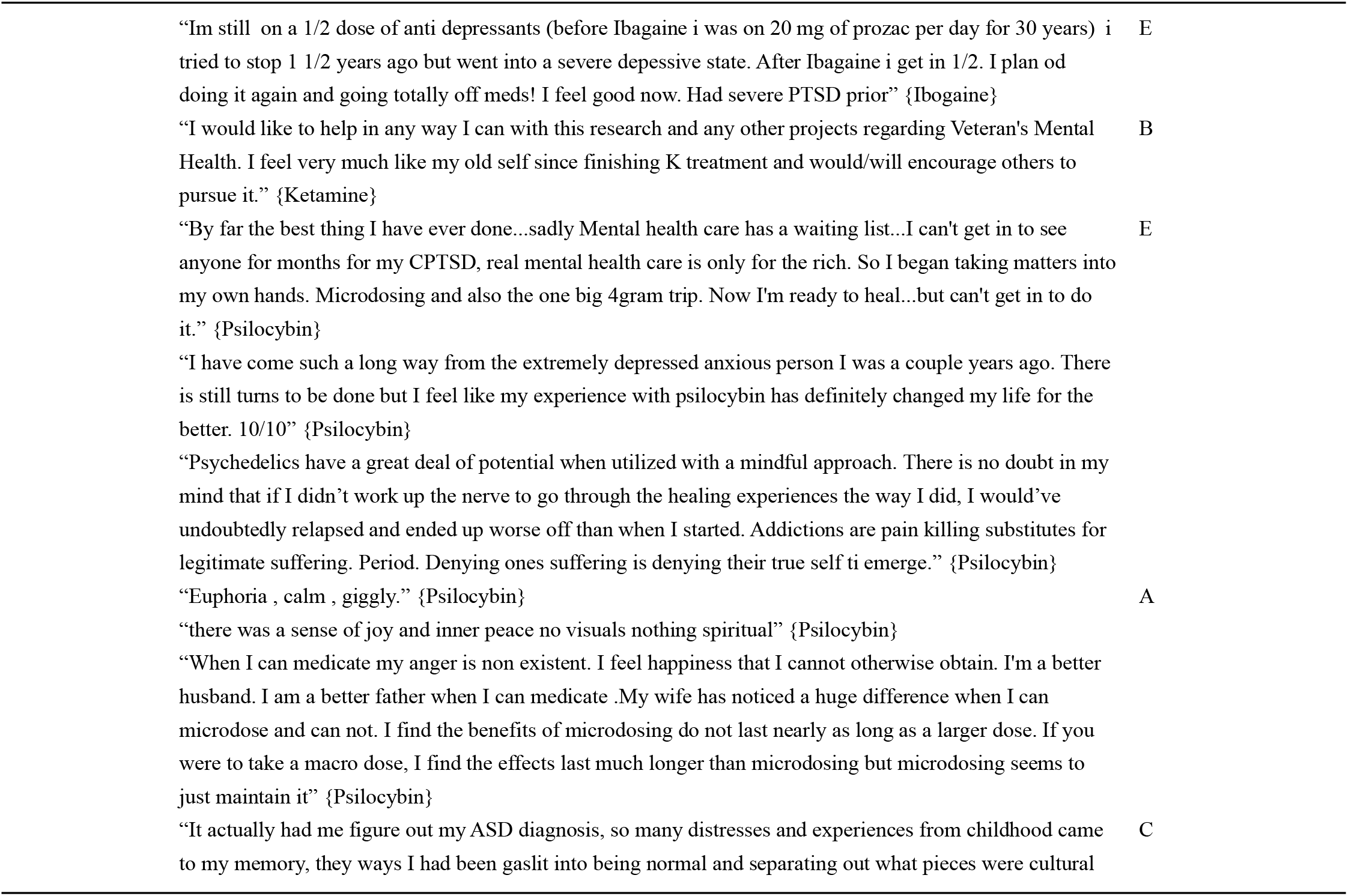
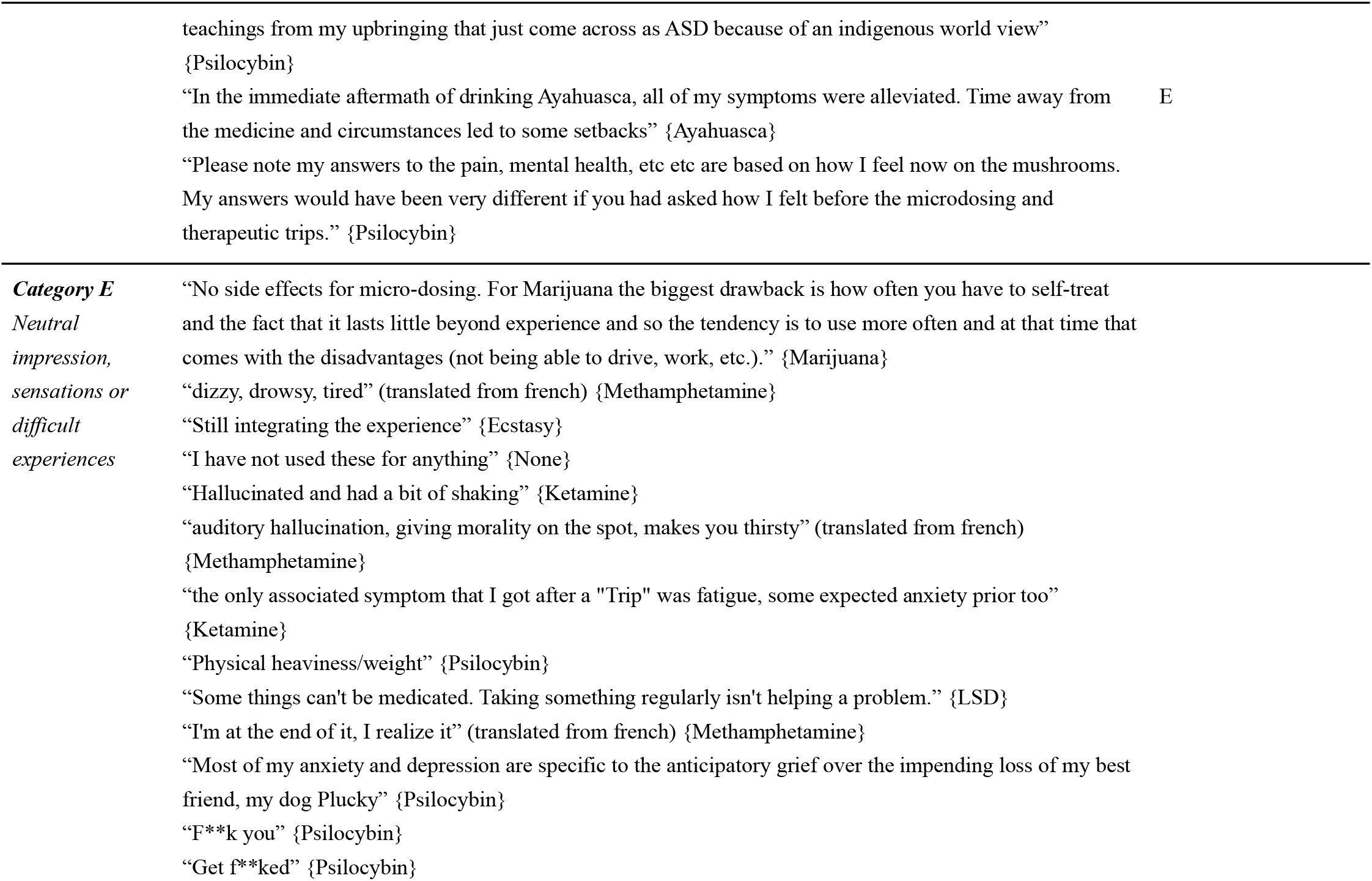
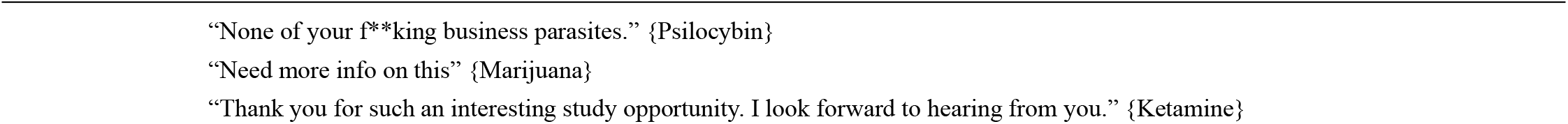

